# Leukocyte telomere length in patients with multiple sclerosis and its association with clinical phenotypes

**DOI:** 10.1101/2020.11.17.20232975

**Authors:** Michael Hecker, Brit Fitzner, Kathrin Jäger, Jan Bühring, Margit Schwartz, Alexander Hartmann, Michael Walter, Uwe Klaus Zettl

**Affiliations:** Rostock University Medical Center, Department of Neurology, Division of Neuroimmunology, Gehlsheimer Str. 20, 18147 Rostock, Germany; Rostock University Medical Center, Institute for Clinical Chemistry and Laboratory Medicine, Ernst-Heydemann-Str. 6, 18057 Rostock, Germany; Charité – Universitätsmedizin Berlin, Institute of Laboratory Medicine, Clinical Chemistry and Pathobiochemistry, Augustenburger Platz 1, 13353 Berlin, Germany

**Author notes:** corresponding authors: Michael Hecker, Michael Walter.

**Keywords:** multiple sclerosis, telomere length, immunosenescence, aging, leukocytes

## Abstract

Aging is a significant factor influencing the course of multiple sclerosis (MS). Accelerated telomere attrition is an indicator of premature biological aging and a potential contributor to various chronic diseases, including neurological disorders. However, there is currently a lack of studies focusing on telomere lengths in patients with MS.

We measured the average leukocyte telomere length (LTL) in biobanked DNA samples of 40 relapsing-remitting MS patients (RRMS), 20 primary progressive MS patients (PPMS) and 60 healthy controls using a multiplex quantitative polymerase chain reaction method. Changes in LTL over a period of >10 years were evaluated in a subset of 10 patients. Association analyses of baseline LTL with the long-term clinical profiles of the patients were performed using inferential statistical tests and regression models adjusted for age and sex.

The cross-sectional analysis revealed that the RRMS group was characterized by a significantly shorter relative LTL, on average, as compared to the PPMS group and controls. Shorter telomeres at baseline were also associated with a higher conversion rate from RRMS to secondary progressive MS (SPMS) in the 10-year follow-up. The LTL decrease over time was similar in RRMS patients and PPMS patients in the longitudinal analysis.

Our data suggest a possible contributory role of accelerated telomere shortening in the pathobiology of MS. The interplay between disease-related immune system alterations, immunosenescence and telomere dynamics deserves further investigation. New insights into the mechanisms of disease might be obtained, e.g., by exploring the distribution of telomere lengths in specific blood cell populations.

**Research in context:** *Evidence before this study:* There is a growing research interest in the relationship between age and the pathophysiology and clinical presentation of multiple sclerosis (MS). Telomere shortening is a hallmark of biological aging. However, the role of telomeres in this chronic immune-mediated neurodegenerative disease has not yet been widely studied. Two research groups provided evidence that the telomeres of immune cells in the peripheral blood are shorter in patients with MS than in healthy subjects.

*Added value of this study:* We found that leukocytes from patients with relapsing-remitting MS (RRMS) are characterized by relatively short telomere lengths (TL). On average, we observed 18% shorter TL in the RRMS patient cohort (n=40) than in the age- and sex-matched healthy control cohort (n=60). We further analyzed the association of TL and long-term clinical outcomes. RRMS patients with shorter TL had a higher rate of converting to secondary progressive MS over a 10-year follow-up period.

*Implications of all the available evidence:* As we and others have shown, TL are generally shorter in MS patients and associated with disease progression, independent of age. These findings suggest a link between biological aging and the heterogeneous clinical course of MS patients. It currently remains unclear whether shortened telomeres in MS are a cause or a consequence of the pathophysiological processes. Further studies with larger patient cohorts and different cell populations will be needed to expand our knowledge of age-related disease mechanisms and the use of TL as a biomarker in MS.

## Introduction

Multiple sclerosis (MS) is a common chronic inflammatory, demyelinating and neurodegenerative disease of the central nervous system (CNS) [1]. The pathological hallmark of MS is the formation of focal lesions in the brain and spinal cord. These lesions are characterized by the infiltration of immune cells, including T cells, B cells and myeloid cells, into the CNS parenchyma, followed by neuroaxonal damage and reactive gliosis [2]. Although the causes of MS are still largely unclear, it has been established that the pathogenic processes are driven by complex interactions of genetic and environmental factors [3]. MS is most often diagnosed in young adults, typically between 20 and 40 years of age, with a two times higher prevalence in women than in men. The clinical presentations and courses of individual patients with MS are very heterogeneous [4]. Most patients (∼90%) suffer from reversible episodes of neurological deficits (also called relapses) lasting several days or weeks during the initial phases of the disease, which are referred to as clinically isolated syndrome (CIS) and relapsing-remitting MS (RRMS) [5]. Within 30 years, the majority of these patients develop a progressive clinical phenotype and permanent physical and cognitive impairments (secondary progressive MS, SPMS) [6]. A subgroup of MS patients (∼10%) shows a gradual worsening of disability without acute exacerbations from disease onset (primary progressive MS, PPMS). MS is currently incurable, but there are more than a dozen approved disease-modifying treatments (DMTs) that can substantially reduce disease activity and slow down progressive neurological deterioration [7-9].

Telomeres play a vital role in preserving genomic stability by protecting the chromosome ends from erosion and end-to-end fusion. In all mammals, telomeres are composed of a (TTAGGG)_n_ tandem repeat DNA sequence (5-15 kb in humans) and a short single-stranded G-rich overhang, which folds back to generate a loop structure governed by proteins that make up the shelterin complex [10,11]. The length of telomeres is known to shorten with each cell division and thus provides insight into the proliferation history of cells. Too short telomeres eventually generate DNA damage signals, resulting in replicative senescence. Telomere attrition is associated with aging but also with a variety of pathological conditions, such as cardiovascular diseases, rheumatoid arthritis, diabetes, cancer and chronic neurological disorders [12,13]. It has been shown that the length of telomeres mediates effects of age on gene expression [14], and some gene regulatory changes have been explained by telomere looping to chromatin [15]. Therefore, it can be assumed that disease mechanisms are affected by telomeres long before the onset of senescence.

There is increasing awareness of the relationship between age and the pathophysiology and clinical manifestation of MS [16]. However, so far little research has been devoted to the roles of telomeres and telomerase, the enzyme that synthesizes telomeric DNA sequences, in this disease. A decreased mRNA expression of the catalytic subunit of telomerase, telomerase reverse transcriptase (TERT), was observed in stimulated peripheral blood mononuclear cells (PBMC) of MS patients as compared to healthy controls [17]. On the other hand, the levels of the telomerase RNA component (TERC) were found to be significantly higher in PBMC-derived CD14+ monocytes of MS patients relative to healthy subjects [18]. Genetic variants at the TERC locus have been associated with both mean leukocyte telomere length (LTL) [19] and risk of developing MS [20]. Moreover, certain lifestyle factors predisposing to MS, such as smoking and obesity [3], are known to correlate with shorter telomeres [21]. Telomere dynamics are also modified by chronic viral infections [22,23]. Recent findings suggest that the etiology of MS is influenced by an interaction between Epstein-Barr virus (EBV) and human herpesvirus (HHV)-6A [24], the latter of which integrates into telomeres of chromosomes [25]. An early study by Hug *et al*. found no significant differences in the length of telomeric DNA in CD4+ and CD8+ T cells obtained from PBMC of RRMS patients and controls [26]. In contrast, Guan *et al*. measured significantly shorter telomeres in the blood of PPMS patients in comparison to RRMS patients, SPMS patients and controls [27]. A later study by this research group provided further evidence that LTL is reduced in MS compared to controls, possibly driven by systemic oxidative stress [28]. More recently, Habib *et al*. reported shortened LTL in all MS subgroups, including RRMS, which is particularly remarkable given the fact that this group was, on average, 9 years and 16 years younger than the controls and the SPMS/PPMS group, respectively [29]. Finally, another study by Krysko *et al*. showed that shorter LTL in MS patients are associated with greater clinical disability as well as brain volume loss over up to 10 years of follow-up [30]. These findings link biological aging with MS phenotypes and motivate further studies on telomeres in patients with MS.

In this study, we used a quantitative polymerase chain reaction (PCR) method to determine relative telomere lengths in blood DNA samples of 120 subjects for comparing relapsing and progressive MS patients with healthy controls. We also analyzed the data in relation to the long-term clinical course of the patients over 10 years. Implications with regard to the use of telomeres as molecular biomarkers and for delineating the pathomechanisms of MS are discussed.

## Materials and methods

### Study cohorts and samples

We used DNA samples that have been stored in our biobank at −20 °C for at least 9 years (sampling period: 12/2003 - 10/2009). The genomic DNA was isolated by standard procedures from peripheral whole blood collected in 8.5 ml PAXgene Blood DNA Tubes (PreAnalytiX). In total, we selected 120 archived samples of 40 RRMS patients, 20 PPMS patients and 60 healthy controls. All 3 groups had a female:male ratio of 1:1 and were well matched for chronological age (patient data given in Table 1, age in years for controls: mean ± standard deviation: 48.1 ± 15.3, median: 51, range: 18-74). From 10 MS patients, a second blood sample was obtained after >10 years from baseline and processed accordingly. DNA quantity and quality were assessed using a NanoDrop 1000 spectrophotometer for calculating DNA concentrations and absorbance ratios at 260/280 nm.

**Table 1:**
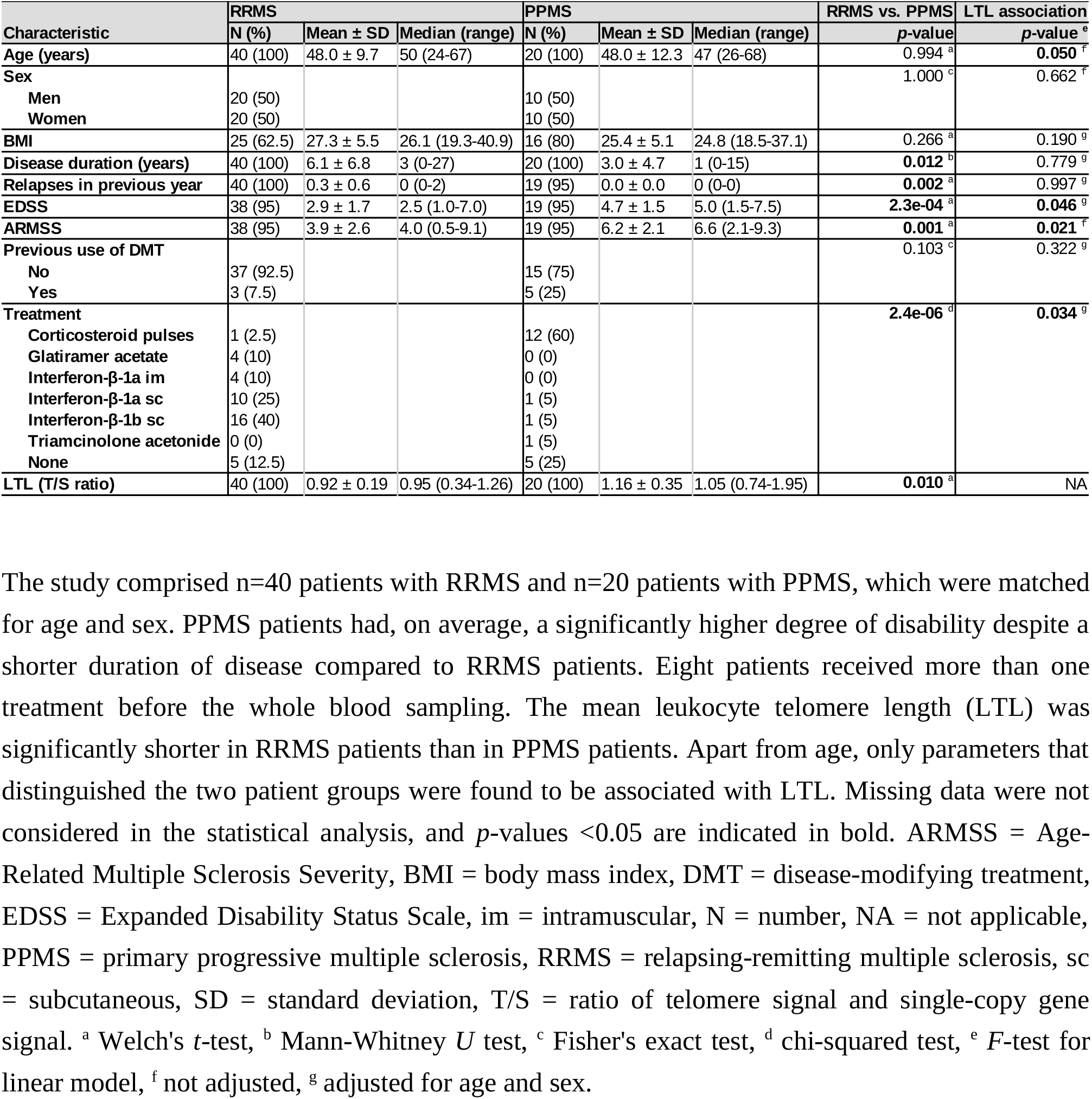
Baseline characteristics of the patients and association with relative telomere length.

Diagnosis of MS has been confirmed according to the McDonald criteria [31,32]. The phenotype of MS was classified on the basis of clinical and paraclinical assessments of relapse occurrence and lesion activity [5]. Functional disability was rated during regular clinical visits using the Expanded Disability Status Scale (EDSS) [33]. Relative severity of disability was evaluated by ranking the baseline EDSS scores based on patients’ age to obtain global Age-Related Multiple Sclerosis Severity (ARMSS) scores [34]. The clinical follow-up period was 10 years. Medical care of the patients always followed routine practice, which means that the patients were treated and monitored at the Department of Neurology of the Rostock University Medical Center according to the guidelines and recommendations of the German Society of Neurology (https://www.dgn.org). The research project has been conducted with the approval by the local ethics committee and in compliance with the principles of the Declaration of Helsinki. All subjects gave prior informed consent for the scientific use of their blood samples. In line with European data protection laws, all records identifying individual subjects are kept confidential.

### Telomere length measurement

We employed the multiplex quantitative PCR telomere length assay published by Cawthon [35], with minor modifications as described elsewhere [36]. Accordingly, telomere (T) signals and single-copy gene (S) signals were measured in comparison to a reference DNA. We used the previously described sequences for the telomere primer pair and the albumin primer pair [35], and the reference DNA sample was generated by pooling all baseline samples. Six concentrations of the reference DNA sample spanning an 243-fold range of DNA concentration were prepared by serial dilution and analyzed in triplicate. The PCR reactions were performed in 384-well plates (Bio-Rad) with Titanium Taq DNA polymerase (Takara Bio) and 20 ng/µl of DNA per sample. SYBR Green fluorescence signals were detected during DNA amplification using a Bio-Rad CFX384 real-time PCR detection system with a C1000 thermal cycler. Melt curve analysis and Cq determination (regression mode) were carried out using the Bio-Rad CFX Manager 3.1 software. Relative quantitation of TL was based on standard curves for the T signal and the S signal. All samples were assayed in 3 runs (at 3 consecutive days). The mean of the 3 T/S values yielded the final relative T/ S ratio as a measure of the average length of the 92 telomeres per cell. Samples with a T/S >1.0 have an average LTL greater than that of the reference DNA. A subset of 30 of the 120 baseline samples was randomly selected for technical replicates (n=15 from MS patients and n=15 from controls). The 20 paired DNA samples from baseline and after >10 years were assayed in an independent experiment following the same protocol and using the same reference DNA. The 170 measurements (including replicates) were conducted in a blinded manner, and thus no sample information (e.g., group assignments) was provided to the experimenter. The full dataset is available in Supplemental Table 1.

### Statistical analysis

All calculations were done in the statistical environment R 3.6.0. Reproducibility across technical replicates and the dependence between LTL and age were assessed by Pearson correlation tests. One-way analysis of variance (ANOVA) was used to examine whether the mean LTL differs significantly between the 3 study cohorts. Sensitivity analyses were performed by removing the most extreme T/S values and by stratifying the subjects by sex. For the comparative analysis of RRMS patients and PPMS patients, we used two-sample two-tailed Welch *t*-tests and Mann-Whitney *U* tests for numerical variables and Fisher exact tests and chi-squared tests for categorical variables. To determine whether clinico-demographic parameters are associated with LTL, we assessed their ability to explain the variance in T/S ratios using *F*-tests for linear models (regression or ANOVA) while adjusting for age and sex as potential confounding variables if appropriate. This was realized with the Anova function of the car package for R [37]. Moreover, univariable and multivariable binary logistic regression models were fitted for the computation of odds ratios (OR) using the trait of having relatively long telomeres (defined by T/S >1) as dependent variable and age, sex and group assignment as independent variables. The change in LTL in the subset of 10 MS patients, for whom samples were available at baseline and after >10 years, was evaluated by paired *t*-test and analyzed for an association with the change in EDSS by linear regression. Kaplan-Meier survival curves were constructed to compare RRMS patients with short (T/S <1) and long (T/S >1) telomeres with regard to the time to the first relapse and the time to SPMS conversion after the blood withdrawal (baseline). Patients who did not experience the respective event and who were lost in the 10-year follow-up period were right-censored. Differences in the time-to-event outcomes were evaluated by hazard ratios (HR) and logrank tests as calculated by Cox regression models using the Breslow method for tie handling. We further assessed the association between LTL and change in disability over 10 years (as rated by the EDSS) in the RRMS cohort using a linear mixed-effects model with a study year by LTL group interaction term [38]. All statistical tests were performed with cases with valid data, and thus cases with missing data were not considered. The significance level was set at α=0.05 without correction for multiple testing as all analyses were exploratory in nature.

## Results

### Patient characteristics

The RRMS patients (n=40) and PPMS patients (n=20) in our study differed with regard to the clinical data (Table 1 and Supplemental Table 1). On average, cases with PPMS presented a much higher degree of disability (as measured by EDSS and ARMSS scores) than cases with RRMS, despite a significantly shorter duration of disease (3.0 ± 4.7 years for the PPMS group vs. 6.1 ± 6.8 years for the RRMS group). In the 12 months prior to the blood sampling, 13 RRMS patients but none of the PPMS patients experienced at least one relapse. The majority of RRMS patients (75%) were treated with interferon-β preparations, whereas the majority of PPMS patients (60%) received corticosteroid pulse therapy, reflecting the standard of care at that time. After 10 years, 26 patients with RRMS and 10 patients with PPMS still had regular visits at our clinic. The remaining patients (n=24) were lost to follow-up, and thus we do not have information on disability progression and relapse activity until the 10-year time point.

### Reproducibility of LTL measurements

The results of the PCR-based telomere length measurements were reproducible across the 3 experimental runs. The inter-assay geometric mean of the coefficient of variation for the T/S ratios over all 150 analyzed blood DNA samples was 5.70%. We also observed a strong agreement in the average T/S ratios of the 30 samples that were measured twice (Pearson correlation: *r*=0.948, *p*=2.0e-15, orthogonal linear regression line: slope=0.947, y-intercept=0.059). For the subsequent analyses, we used the mean T/S of these technical duplicates. The average T/S values (that is, the relative LTL) of the 120 independent samples (n=60 from German MS patients and n=60 from German controls) ranged from 0.34 to 2.30 (mean ± standard deviation: 1.06 ± 0.32).

### Comparison of telomere lengths in MS and controls

We observed significantly shorter telomeres in the group of RRMS patients as compared to PPMS patients and healthy subjects (one-way ANOVA *p*=0.003) (Figure 1A). The difference between RRMS cases and controls was robust in the post-hoc sensitivity analyses where we excluded the minimum and/or maximum value and where we analyzed females and males separately (*t*-test *p-* values <0.05). The patients with relapsing MS also showed a much lower odds of having relatively long telomeres (T/S >1) in the multivariable binary logistic regression analysis (OR=0.399, 95% confidence interval: 0.170-0.913) (Figure 1B), whereas no significant differences were found between the PPMS group and controls as well as between women and men (*t*-test *p*=0.768). As expected, the age of the 120 subjects was negatively associated with LTL (Pearson correlation: *r*=-0.222, *p*=0.015). This trend was seen independently in all 3 study cohorts as well as in both sexes (Pearson correlation: *r* <-0.128, univariable logistic regression: OR <0.984), despite considerable interindividual variation in the data (Figure 1C).

**Fig. 1:**
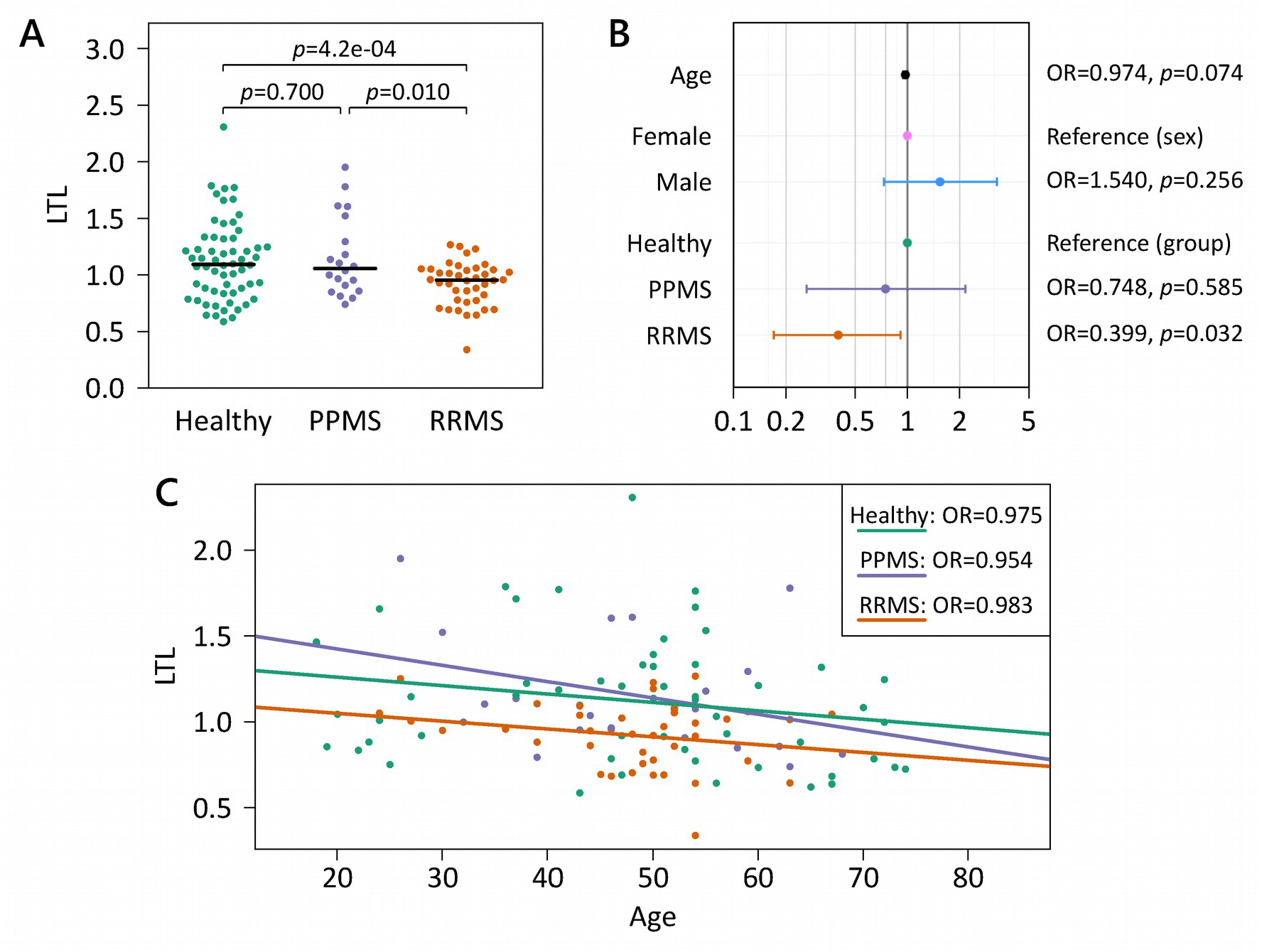
Relationship between telomere length and age, sex and diagnosis of MS. Average leukocyte telomere lengths (LTL) were measured in age- and sex-matched healthy subjects (n=60), RRMS patients (n=40) and PPMS patients (n=20). Using the quantitative polymerase chain reaction method by Cawthon [35], telomere (T) signals and single-copy gene (S) signals were measured in comparison to reference DNA, yielding relative T/S ratios. (**A**) Beeswarm plot showing differences in LTL between the groups (one-way analysis of variance *p*=0.003). Two-tailed Welch *t*-test *p*-values are reported above the brackets. The black lines indicate the medians per group (1.09 for healthy controls). (**B**) Associations with relatively long telomeres (T/S >1). Visualized are the odds ratios (OR) and 95% confidence intervals (CI) from the multivariable binary logistic regression analysis. The OR for age is given for each one-year increase (95% CI 0.944-1.002). (**C**) High age correlates with short telomeres. Linear regression lines are shown for each group. MS = multiple sclerosis, PPMS = primary progressive multiple sclerosis, RRMS = relapsing-remitting multiple sclerosis.

### Relationships between LTL and clinical features

We checked whether there are associations between the obtained T/S ratios and the clinical parameters of the 60 MS patients. EDSS, ARMSS and treatment at the time of assessment (baseline) were significantly related to the relative telomere lengths (*F*-test *p*-values <0.05). However, these are presumably indirect associations, reflecting the apparent differences between RRMS patients and PPMS patients (Table 1). Accordingly, these associations disappeared when the patients were stratified by the course of MS (*p*-values >0.31). The only independent variable that had an influence on LTL in this analysis was again age (*F*-test *p*=0.050).

### Long-term clinical course in relation to telomere length

Telomere shortening was seen for each of the 10 MS patients with LTL measured at two time points (annualized LTL decrease: mean ± standard deviation: 0.014 ± 0.008, median: 0.013, range: 0.003-0.026) (Figure 2A). The LTL decrease was significant for both RRMS patients (n=6) and PPMS patients (n=4) (paired *t*-test *p*-values <0.05). Change in LTL was weakly associated with change in EDSS in linear regression. For every 0.01 annualized LTL decrease, the 10-year increase in EDSS was estimated to be 0.29 (*F*-test *p*=0.182). However, the cohort with longitudinal blood sampling was too small to draw meaningful conclusions in this respect.

**Fig. 2:**
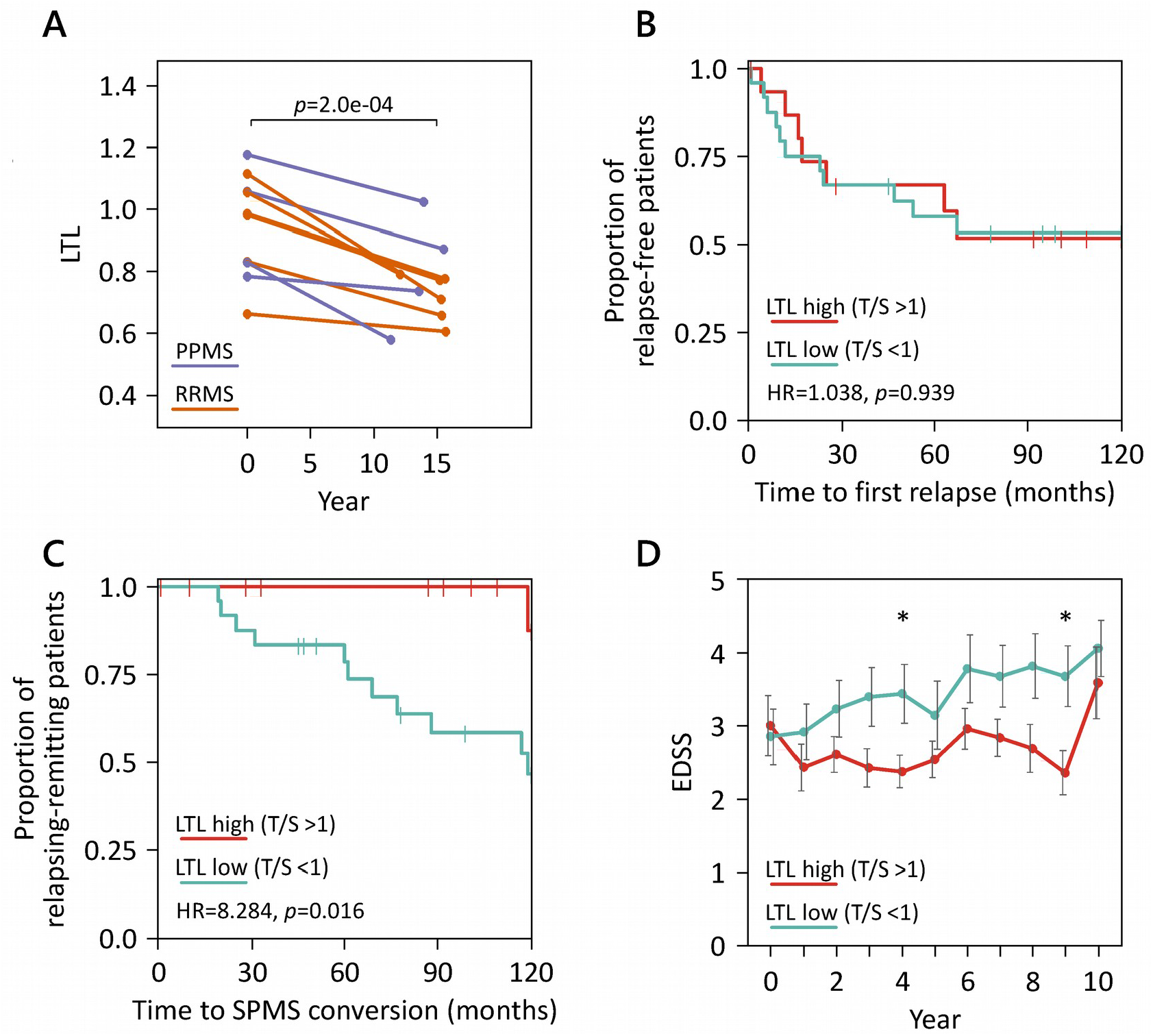
Telomere dynamics and long-term clinical course stratified by telomere length. **(A)** For 10 MS patients, relative LTL was measured at baseline and after >10 years. The connected scatterplot shows similar LTL decrease over time (paired *t*-test *p*=2.0e-04), independent of disease subtype at baseline. (**B**-**D**) The group of RRMS patients was split into two groups with low (n=24) and high (n=16) LTL, respectively. They were followed up for 10 years after the blood sampling. **(B)** Kaplan-Meier curves for relapse-free survival. Overall, the curves of both groups were similar. The Cox proportional hazard ratio (HR) and the logrank test *p*-value are given in the figure, and right-censored observations are indicated by vertical dashes. (**C**) Kaplan-Meier plot showing that RRMS patients with low LTL have a much higher risk for conversion to SPMS. (**D**) Comparison of disability progression for RRMS patients with low and high LTL at baseline. There was a tendency of elevated average EDSS scores in the group of patients with relatively short telomeres. Only patients with available data at the applicable time points were included in this analysis. Asterisks (*) denote significant differences (two-tailed Welch *t*-test *p*-values <0.05). Error bars indicate standard errors. EDSS = Expanded Disability Status Scale, LTL = leukocyte telomere length, MS = multiple sclerosis, PPMS = primary progressive multiple sclerosis, RRMS = relapsing-remitting multiple sclerosis, SPMS = secondary progressive multiple sclerosis, T/S = ratio of telomere signal and single-copy gene signal.

For the subgroup of RRMS patients, we analyzed whether the LTL at baseline may have a predictive value with regard to disease progression over 10 years. The time to the first documented relapse was similar in those patients with short telomeres (n=24, T/S <1, LTL low) and those patients with long telomeres (n=16, T/S >1, LTL high) (Figure 2B). However, the proportion of patients that converted to SPMS was significantly different (HR=8.284, logrank test *p*=0.016, with adjustment for age: HR=8.308, *p*=0.050). At the end of the 10-year follow-up period, 11 patients of the LTL low group but only 1 patient of the LTL high group were classified as SPMS (Figure 2C).

Patients with relatively short telomeres at baseline also showed a continued worsening of disability over time and had nominally higher EDSS scores than LTL high patients on average (Figure 2D). A linear mixed-effects model analysis revealed a significant main effect for study year (*F*-test *p*=2.2e-18) but no significant interaction effect with telomere length (*p*=0.277). Thus, no clear association between LTL and the rate of change in EDSS could be found in the present data, although individuals with longer telomeres tended to have a more favorable course of disease.

## Discussion

Telomere shortening during aging is possibly connected to pathological and immunological processes in MS. In our PCR-based cross-sectional analysis, we detected significantly shorter LTL in RRMS patients than in healthy controls. This difference was evident for both men and women. Moreover, RRMS patients with relatively high LTL were less likely to convert to SPMS within 10 years. This suggests that age-related disease mechanisms may be captured by LTL as a biomarker for immunosenescence. Our study adds evidence to the literature that individual diversity in biological aging may contribute to the heterogeneity in the course of MS.

Reduced average telomere lengths in blood cells of patients with MS were previously reported by two research groups [27-29]. In accordance with our study, significantly lower LTL in RRMS compared to controls were recently described by Habib *et al*. [29]. However, Guan *et al*. found that specifically PPMS patients have much shorter telomeres [27], which we could not confirm based on our data. This discrepancy might be explained by the fact that quite different study populations were used. Their RRMS group differed from ours, e.g., in terms of sample size (n=19 vs. n=40), genetic background (Asian vs. Western European), average age (33.8 years vs. 48.0 years) and disease severity (benign cases with EDSS <3.0 vs. all cases). Moreover, Guan *et al*. used Southern blots for the measurement of telomere lengths [27,28]. In contrast to quantitative PCR assays, this method also detects a variable portion of subtelomeric DNA, dependent on the restriction enzymes used [39,40]. The so far only other telomere study that compared MS patients with healthy subjects was confined to T cells [26]. In this study, no significant differences were observed. It is important to note that shorter telomeres in the mixture of leukocytes may not necessarily be reflected in the T cell population of MS patients. Although telomere lengths are positively correlated within an individual among T cells, B cells and monocytes, the rates of change differ between the cell types [41,42]. The decline in telomere length with age is also more pronounced in lymphocytes than in granulocytes [43]. Therefore, studies investigating telomeres in specific immune cell subtypes of MS patients are clearly needed.

Accelerated somatic telomere length shortening during aging is presumably part of the altered immune response in MS and may contribute to neurodegeneration. However, it remains unclear whether there is a direct role of telomeres in the pathogenesis of MS or whether shortened telomeres are merely a surrogate marker of increased oxidative stress and/or cell turnover. Inflamm-aging and immunosenescence are closely related to each other and are associated with a variety of immune function changes, starting with the involution of the thymus [44,45]. The average LTL assessed in our study reflects the proliferative potential and chromosomal stability of the circulating cells of the immune system [11]. The level of immunocompetence critically depends on cell renewal and clonal expansion of T- and B-cell populations [46]. Telomere attrition in these cells ultimately triggers a persistent DNA damage response (DDR) that leads to cellular senescence or apoptosis [47]. As a consequence, the ability to initiate and sustain robust adaptive immune responses deteriorates while the number of naive T and B cells in the peripheral blood decreases with age [46]. On the other hand, infection with cytomegalovirus (HHV-5) has been strongly implicated in premature immunosenescence by causing expansion of late-differentiated CD8+CD28-T cells [22,48,49]. Progressive shortening of telomeres also provokes telomere looping changes, which has widespread effects on chromatin organization and gene regulation long before the initiation of a DNA damage signal [15]. The telomeric integration of HHV-6 may affect the expression of human genes by interfering with this phenomenon [50].

It can be speculated that disease-specific defects in telomere biology may promote the disruption of telomeric structures. Therefore, additional studies on the expression of telomere maintenance genes and LTL-regulated genes would be useful to explore potential mechanisms underlying a premature loss of telomere repeats in MS. Further investigations are also required to better understand how genetic, epigenetic and nongenetic determinants of telomere length may interact prior to disease onset and in the course of MS. Lifestyle and environmental factors that have been associated with dysfunctional telomeres and that are actively researched in the context of MS include smoking, excessive dietary intake, lack of physical activity, chronic psychological and oxidative stress, chronic inflammation and exposure to certain persistent viral infections [21,22,51-53]. The particular vulnerability of telomeric sequences to oxidative damage [54] may form the underlying basis of risk factors of accelerated telomere attrition.

While we analyzed a well-characterized cohort of MS patients, this study also has some limitations. As our analyses were exploratory, any significant result reflects a statistical relationship subject to further testing for independent confirmation. On the other hand, given the substantial heterogeneity in individual telomere length, our study was underpowered to detect moderate effects. For instance, a negative association of LTL and BMI, which has been repeatedly reported in the literature [51], was noted in the data but did not reach significance (*F*-test *p*=0.190). In line with the findings by Krysko *et al*. [30], we did not observe an association between LTL and DMT at baseline in the RRMS group (*p*=0.713). However, more potent therapies for MS have been approved in the past years [8]. As some of them induce profound changes in the immune repertoire, they may influence markers of immunosenescence as well. In our study, RRMS patients with relatively short telomeres had a higher risk of conversion to SPMS in the following 10 years. This inverse relationship was also reported by Krysko *et al*. [30], although it was not significant in their study. Larger cohorts are necessary to verify this observation. Another issue is that progression of MS overlaps with aging-related deficits [16] and that we did not capture other potential confounders, such as smoke exposure and comorbidities. Therefore, it is difficult to disentangle direct and indirect relationships between telomere length and clinico-demographic parameters. It also remains unclear whether a lower LTL in RRMS patients may be explained by reduced telomerase activity, which is tightly regulated during lymphocyte differentiation [41,46]. Another open question is how well mean LTL reflects telomere length in the CNS tissue of MS patients. In general, telomere length is highly correlated among tissue types [14], but immunological and CNS-specific aging may play independent roles in the course of MS.

Further studies employing telomere-based and/or epigenetic measures of biological age [55,56] are warranted in the field of MS. More labor-intensive methods can provide information on the shortest (not just average) telomere lengths and on telomeres at individual chromosome arms [57]. Furthermore, new insights into the pathobiology of MS might be gained by investigating the transcription of damage-induced non-coding RNAs and the recruitment of DDR proteins to sites of DNA damage in telomere-deprotected cells [58]. Deeper knowledge on the molecular consequences of short telomeres in specific cell subsets may yield novel biomarkers of disease activity and disability progression. It can be also speculated that behavioral interventions that may influence telomere dynamics (e.g., smoking avoidance, stress reduction, dietary change and physical activity) [52,59,60] can modulate immune cell functions in MS patients. Eventually, targeting aging-related mechanisms could be an integral part of patient care in the near future [61,62].

To conclude, our study supports a link between telomeres and MS. We observed significantly lower LTL in subjects with RRMS. In those patients, short telomeres at baseline were associated with a higher rate of conversion to SPMS and sustained worsening of disability in the long-term follow-up. This suggests that biological aging contributes to the heterogeneous clinical presentation of MS patients, which needs to be elaborated in more detail by subsequent studies. Further research is also required to explore the molecular events involved in telomere attrition in individual immune cell populations. This will yield a better understanding of the possible role of telomeres in the inflammatory and neurodegenerative disease processes underlying MS.

## Data Availability

All relevant data of this study are provided in Supplemental Table 1.

## Abbreviations

ANOVA: analysis of variance
ARMSS: Age-Related Multiple Sclerosis Severity
BMI: body mass index
CI: confidence interval
CIS: clinically isolated syndrome
CNS: central nervous system
DDR: DNA damage response
DMT: disease-modifying treatment
EBV: Epstein-Barr virus
EDSS: Expanded Disability Status Scale
HHV: human herpesvirus
HR: hazard ratio
im: intramuscular
LTL: leukocyte telomere length
MS: multiple sclerosis
N: number
NA: not applicable
OR: odds ratio
PBMC: peripheral blood mononuclear cells
PCR: polymerase chain reaction
PPMS: primary progressive multiple sclerosis
RRMS: relapsing-remitting multiple sclerosis
S: single-copy gene signal
sc: subcutaneous
SD: standard deviation
SPMS: secondary progressive multiple sclerosis
T: telomere signal
TERC: telomerase RNA component
TERT: telomerase reverse transcriptase

## Authors’ contributions

MH designed the study, wrote the manuscript and analyzed the data. BF prepared the DNA samples for the analyses. KJ performed the laboratory experiments. JB compiled the relevant literature and participated in the analysis and interpretation of the data. MS was responsible for blood sampling and clinical documentation. AH assisted in the documentation of the experimental procedures. MW coordinated and supervised the telomere length measurements and contributed to the Discussion section. UKZ participated in the study design and provided important intellectual content. The authors had full access to the data of this study. All authors improved the drafts and approved the final version of the manuscript.

## Acknowledgments

We thank Christa Tiffert and Antje Bombor for coordinating patient care. We are grateful to Nele Retzlaff and Deborah Sonnenberg for clinical documentation. We thank Paul Blaschke, Frank Blaschke and Edgar Piontek for recruiting control subjects. We thank Dirk Koczan and Ildikó Tóth for laboratory assistance.

## Data sharing statement

All relevant data of this study are provided in Supplemental Table 1.

